# Joint Effects of Real-World Cue Exposure and Affective States on Momentary Alcohol Craving in Adults with Alcohol Use Disorder

**DOI:** 10.64898/2026.05.18.26353518

**Authors:** Abhishek Aggarwal, Peter M. Monti, Kittichai Promrat, Molly Magill, Jessica L. Mellinger, Hayley Treloar Padovano

**Affiliations:** Center for Addiction and Disease Risk Exacerbation, Brown University, 121 S. Main Street, Providence, RI 02912, United States; Center for Alcohol and Addiction Studies, Brown University, 121 S. Main Street, Providence, RI 02912, United States; Brown University Health, Providence, RI, United States; Henry Ford Health, Detroit, MI, United States; Michigan State University, Lansing, MI, United States

**Keywords:** alcohol use disorder, ecological momentary assessment, cue reactivity, craving, affect

## Abstract

**Background:** Alcohol use disorder (AUD) is marked by high relapse rates often driven by craving, yet less is known about whether *in vivo*, social, and place-based alcohol cues are differentially associated with craving across affective states. This study examined independent and affect-contingent associations of these cues with momentary craving in adults with AUD enrolled in an alcohol intervention study.

**Methods:** Thirty-three adults with AUD completed up to four daily ecological momentary assessments (EMA) for 28 days. EMA prompts assessed craving, *in vivo* alcohol exposure, being around usual drinking partners, being in usual drinking places, and high-arousal positive affect (PA) and negative affect (NA). Multilevel mixed-effects models adjusted for demographics, intervention phase (1 = post, 0 = pre), AUD severity, and temporal and contextual covariates.

**Results:** EMA compliance was high (*Median* per-participant = 86.6%). Within-person elevations in *in vivo* alcohol exposure and being around usual drinking partners were independently associated with greater momentary craving, whereas being in usual drinking places was not. *In vivo* alcohol exposure was more strongly associated with craving during higher-than-usual PA (*β* = 0.08, *p* = .032), whereas being in usual drinking places was more strongly associated with craving during higher-than-usual NA (*β* = 0.06, *p* = .036), adjusting for intervention phase (which was associated with lower craving).

**Conclusions:** Findings support the need for personalized just-in-time adaptive interventions tailored to high-risk, momentary cue–affect contexts in AUD, beyond low-frequency clinician-delivered feedback that may reduce average craving but not fully address real-time risk. ClinicalTrials.gov registration: NCT05135767.

## 1. Introduction

Alcohol use disorder (AUD) remains one of the most prevalent psychiatric conditions worldwide. In the United States, an estimated 28.9 million individuals aged 12 and older (10.2%) met past-year criteria for AUD in 2023 (Substance Abuse and Mental Health Services Administration, 2024). Despite the magnitude of the disorder, fewer than one in five adults with lifetime AUD ever access treatment, and among those who do, 60–70% return to heavy drinking within three months post-discharge (Carvalho et al., 2019; MacKillop et al., 2022). Craving, formally incorporated as a DSM-5 diagnostic criterion (American Psychiatric Association, 2013), is increasingly recognized as a central clinical mechanism in the maintenance of AUD (Sayette, 2016). A recent meta-analysis of 237 studies found that cue and craving indicators were prospectively associated with more than double the odds of subsequent substance use or relapse (Vafaie and Kober, 2022). Cue reactivity, defined as the subjective, physiological, and neural responses elicited by alcohol-associated stimuli, is therefore widely regarded as the proximal mechanism through which everyday environmental triggers translate into lapse and relapse (Carter and Tiffany, 1999).

According to the incentive sensitization theory (IST), repeated alcohol use sensitizes the brain’s mesocorticolimbic dopamine systems, causing them to attribute excessive “incentive salience” to alcohol-related cues (Berridge and Robinson, 2016; Robinson and Berridge, 1993, 2025). Critically, IST distinguishes between the sensitized “wanting” system (incentive motivation) and the “liking” system (hedonic impact). In AUD, environmental cues can trigger an intense, pathological “wanting” that persists even if the actual pleasure derived from drinking has diminished (Robinson and Berridge, 2025). As these cues are learned through repeated alcohol use in specific social and physical contexts, understanding cue reactivity in AUD requires assessing the varied real-world cues that individuals encounter in daily life. In this regard, ecological momentary assessment (EMA) studies have identified three broad cue domains: (1) *in vivo* cues, i.e., direct exposure to alcohol (Kohen et al., 2023; Treloar Padovano and Miranda, 2021; Wycoff et al., 2023); (2) personalized social cues such as being around peers or partners with whom one typically drinks (Acuff et al., 2024; Fairlie et al., 2025; Meisel et al., 2023); and (3) personally meaningful place-based cues (e.g., bars, restaurants) (Fatseas et al., 2015; Kuerbis et al., 2020; Serre et al., 2024). EMA has therefore become a key method for capturing cue exposure and reactivity as they occur in daily life, with the goal of informing personalized interventions for AUD (Serre et al., 2015; Trull and Ebner-Priemer, 2013, 2020).

Complementary affective-processing models of negative reinforcement suggest that avoidance of negative affect can also trigger craving when drinking has repeatedly been paired with alcohol’s temporary relief effects (Baker et al., 2004). Consistent with this view, laboratory studies show that experimentally induced NA reliably increases alcohol craving (Bresin et al., 2018), and EMA studies indicate that within-person increases in NA predict greater momentary craving, particularly among individuals with stronger coping motives (Waddell et al., 2021; Wycoff et al., 2021). Evidence linking NA to actual drinking in daily life, however, is less consistent. A recent individual-participant-data meta-analysis of 69 EMA studies (N = 12,394) found that individuals were not more likely to drink on high-NA days, but were more likely to drink, and to drink heavily, on high-positive-affect (PA) days (Dora et al., 2023). Notably, that meta-analysis aggregated mood scores into daily averages, which may obscure within-day affect–craving dynamics. This pattern is nonetheless consistent with motivational models that distinguish negative-reinforcement drinking from positive-reinforcement or enhancement-based drinking pathways (Cooper, 1994). From this perspective, high-arousal PA (e.g., feeling energetic, excited) plausibly engages approach motivation and incentive salience circuitry, and may therefore facilitate cue-elicited “wanting” (Berridge and Robinson, 2016; Cofresí et al., 2019). However, PA has been markedly understudied in the craving literature relative to NA.

Although alcohol cues and affective states have often been studied as parallel contributors to craving, theoretical and empirical work increasingly suggests that they may operate jointly in daily life. Dynamic models of relapse conceptualize cue reactivity as a momentary process that varies across emotional and contextual states (Witkiewitz and Marlatt, 2004). Affective states may shape craving by altering attention, reward sensitivity, and motivational processing, thereby increasing the salience and incentive value of alcohol cues when affect is heightened (Sinha, 2013). Consistent with this view, laboratory studies show that alcohol cues paired with negative affect or stress elicit stronger cravings and prospectively predict relapse risk (Coffey et al., 2010; Cooney et al., 1997; Sinha et al., 2011). Daily-life evidence for interactions between cue exposure and affect is more recent but increasingly direct. In a 14-day daily-diary study of non-treatment-seeking adults with AUD, NA strengthened the cue–drinking association while PA reduced it (Meredith et al., 2025). A companion daily-diary study during early abstinence further reported synergistic effects of mood and cue exposure on next-day craving (Baskerville et al., 2025).

Despite this growing literature, two key gaps limit understanding of how alcohol cues and affect jointly shape momentary craving in adults with AUD. First, EMA cue-reactivity studies have largely focused on adolescents, emerging adults, or non-treatment-seeking heavy drinkers, with fewer studies examining adults with AUD (Cofresí et al., 2025; Kang et al., 2022; Meisel et al., 2023). Second, existing studies have either examined cue-reactivity for each cue domain separately, used generic indicators (e.g., any drinking-related location) rather than idiographic, person-specific cues, or relied on a single composite cue indicator (Baskerville et al., 2025; Kuerbis et al., 2020; Meredith et al., 2025), leaving unclear which cue types uniquely predict craving when modeled together (Robinson and Berridge, 2025).

The present study addresses these gaps using smartphone-based EMA data from adults with AUD enrolled in an alcohol intervention study. First, we examined three real-world alcohol cues simultaneously in multilevel models, namely an *in vivo* cue (direct alcohol exposure), a personalized social cue (being around people with whom one usually drinks), and a personalized place cue (being in places where one usually drinks), hypothesizing that each would be independently associated with greater momentary craving. Second, using separate moderation models, we tested the hypothesis that high-arousal positive affect (PA; happy, energetic) and negative affect (NA; anxious, stressed) would amplify the within-person associations between cue exposure and craving. This knowledge is clinically consequential because cue–affect combinations may reveal discrete moments when craving is most likely to escalate and when just-in-time interventions delivering coping, distress-tolerance, or reward-regulation support may be most needed (Nahum-Shani et al., 2018).

## 2. Methods

### 2.1 Participants and Procedures

The analytic sample comprised 33 adults aged 21 or older (*mean* age = 46.0 years, *SD* = 12.55; 51.5% male; 30.3% Hispanic) with AUD (mean AUDIT total = 17.09, *SD* = 8.59). To qualify for participation, individuals were required to have reported alcohol intake exceeding risk thresholds (≥14 drinks/week for men or ≥7 drinks/week for women) during any 7-day period within the 90 days prior to enrollment. The entire intervention study, including EMA, was offered in English and Spanish, increasing rigor and research representation through including monolingual Spanish speakers. The EMA protocol was a part of a nonrandomized parallel-assignment intervention study designed to increase awareness of alcohol’s effects on liver health among participants with AUD, including a subset with diagnosed alcohol-associated liver disease. Pre-registration of the study protocol is available publicly at ClinicalTrials.gov (NCT05135767). All procedures received approval from the Brown University Institutional Review Board (IRB; 2011002840), with Lifespan IRB (1712272) as a relying site. All participants provided written informed consent prior to enrollment.

The EMA data spanned a total observation window of 28 calendar days per participant, beginning with a preintervention baseline phase lasting up to 10 calendar days. After the baseline phase, participants received personalized liver-health and drinking-precursor feedback via a 60-minute motivational interviewing session and a 30-minute booster session scheduled after participants completed the full 28-day EMA reporting period. Participants completed up to 4 EMA reports per day to capture drinking patterns and precursors both before and after the intervention. Audible notifications prompted reports within four blocked intervals to ensure a representative sample across waking hours (8:00 a.m.–11:30 a.m., 11:31 a.m.–3:00 p.m., 3:01 p.m.–6:30 p.m., and 6:31 p.m.–10:00 p.m.). Reports not completed within 30 minutes were marked as missed. To maximize compliance, participants received up to three reminders per prompt via push notification, text message, and automated phone call. The final analytic sample comprised 2,369 EMA prompts from 33 participants after excluding EMA reports completed during active drinking (see *Measures: Momentary current drinking*).

### 2.2 Measures

#### 2.2.1. Momentary craving

At each prompt, participants reported their current alcohol craving on an 11-point sliding scale (0 = *Not at all*, 10 = *Extremely*) in response to the item, *“How strong is your CRAVING to drink ALCOHOL RIGHT NOW?”* (Sayette, 2016; Serre et al., 2015; Toulami et al., 2025).

#### 2.2.2. Real-world alcohol cues

Three real-world cue indicators were derived from prompt-level responses. The *in vivo* cue was assessed with “Can you SEE ALCOHOL RIGHT NOW?” Endorsement of “Yes, in real life [bottle, glass]” was coded 1, and all other responses, including “No,” “Yes, on TV, ad, online,” and “Yes, saw liquor store, bar, other drinking place,” were coded 0. Personalized social and place cues were derived from multiple-response items assessing whether participants were “with PEOPLE I usually drink with” or “in a PLACE where I usually drink.” Endorsement of each item was coded 1 for the corresponding cue and 0 otherwise.

#### 2.2.3. Momentary current drinking

At each prompt, participants reported whether they were currently drinking via a single binary item: *“Are you DRINKING alcohol RIGHT NOW?”* (0 = *No*, 1 = *Yes*).

#### 2.2.4. Affect composites

Each prompt assessed momentary affective states on 0–10 sliding scales (0 = *Not at all*, 10 = *Extremely*) in response to four items asking how *happy*, *energetic*, *anxious*, and *stressed* the participant felt (e.g., *“How HAPPY do you feel RIGHT NOW?”*). High-arousal PA was computed as the sum of happy and energetic (range 0–20), and high-arousal NA was computed as the sum of anxious and stressed (range 0–20), consistent with prior EMA work conceptualizing PA and NA along arousal–valence dimensions (Russell, 1980; Watson et al., 1988).

#### 2.2.5. Time of day

The time at which the prompt was submitted was categorized into three windows: Morning (before 11:30; reference category), Afternoon (11:30–14:59), and Evening (15:00 onward).

#### 2.2.6. Covariates

All multilevel models adjusted for participant age, gender (1 = male, 0 = female), education (last grade completed), Hispanic ethnicity, financial status, baseline AUDIT total score (centered), intervention phase (1 = post, 0 = pre), time of day (afternoon and evening dummy indicators with morning as reference), weekend vs weekday, and between-person levels of all cues and affect.

### 2.3 Statistical Analysis

EMA reports completed while participants reported currently drinking (*n =* 369, 13.5%) were excluded before analysis so that cue–craving associations reflected responses to environmental cues rather than the effects of alcohol already consumed. This decision was supported by prior EMA evidence showing that recent alcohol use moderates cue–craving associations, with cue-elicited craving differing between non-drinking and post-drinking moments (Kohen et al., 2023). All time-varying predictors were partitioned into within-person (deviations from each participant’s mean) and between-person (each participant’s mean centered around the grand mean) components (Curran and Bauer, 2011; Wang and Maxwell, 2015). Multilevel linear mixed-effects models with random intercepts for participants were estimated using the lmer function from the lme4 R package (Bates et al., 2015), with degrees of freedom approximated via Satterthwaite’s method as implemented in lmerTest (Kuznetsova et al., 2017). Three primary models were fit. Table 3 reports a model regressing momentary craving on all three real-world cues simultaneously. Tables 4 and 5 examined the moderating role of momentary affect in the momentary association between each cue and craving. The random-slope structure was selected via systematic likelihood-ratio testing; the resulting structure was strongly favored over random-intercept-only specifications (all *χ²* > 145, *df* = 5, *p* < .001), indicating substantial between-person heterogeneity in cue and affect reactivity. Model convergence was achieved using the bobyqa optimizer (Powell, 2009). Analyses were conducted in R version 4.3.3 (R Core Team, 2024).

## 3. Results

### 3.1 Sample and EMA Compliance

Descriptive statistics for the analytic sample are presented in Table 1. Across the 33 participants, the mean age was 46.0 years (*SD* = 12.55); 51.5% identified as male, 30.3% as Hispanic, and the average baseline AUDIT total score was 17.09 (*SD* = 8.59). Participants reported *in vivo* alcohol exposure at 15.4% of moments, being around people with whom they usually drink at 15.4%, and being in places where they usually consume alcohol at 41.0%. Compliance to the EMA protocol was computed prior to removing moments in which participants reported drinking (*n* = 369, 13.5%). Across the 33 participants, 2,738 of 3,556 expected prompts were completed, yielding an overall compliance rate of 77.0% (*mean* per-participant = 76.6%; *median* = 86.6%).

**Table 1.**
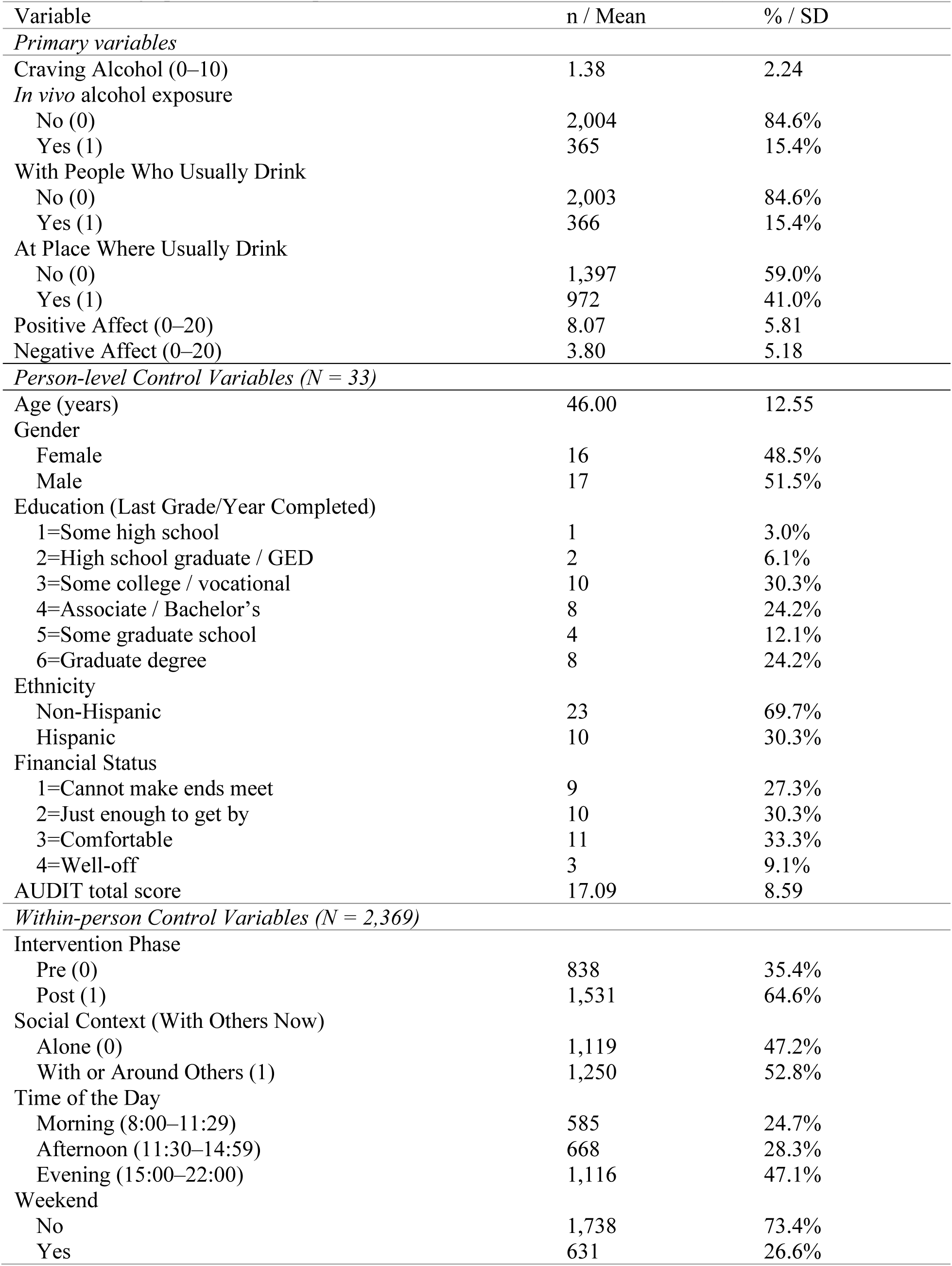
Demographics and Descriptive Statistics (N = 33; Observations = 2,369)

Table 2 displays bivariate correlations among the primary variables, with within-person correlations below the diagonal, between-person correlations above, and intraclass correlation coefficients (ICCs) on the diagonal. ICCs indicated substantial within-person variation in all variables, with ICCs ranging from .18 (*in vivo* alcohol exposure) to .75 (PA). As these bivariate correlations do not adjust for the nested structure of the data, they are presented for descriptive purposes only.

**Table 2.** Correlations and Intraclass Correlation Coefficients for Main Variables (within-person N = 2,369; between-person N = 33)

|  | 1 | 2 | 3 | 4 | 5 | 6 |
| --- | --- | --- | --- | --- | --- | --- |
| 1. Craving | *0.46* | 0.17 | 0.08 | 0.24 | -0.30† | 0.60*** |
| 2. *In vivo* alcohol exposure | 0.07*** | *0.18* | -0.05 | 0.38* | -0.16 | 0.27 |
| 3. People Usually Drink | 0.08*** | 0.18*** | *0.20* | 0.41* | -0.25 | 0.01 |
| 4. Place Usually Drink | 0.03 | 0.20*** | 0.20*** | *0.38* | -0.29 | 0.16 |
| 5. Positive Affect | 0.07** | 0.02 | 0.04† | -0.11*** | *0.75* | -0.48** |
| 6. Negative Affect | 0.08*** | -0.01 | -0.04† | -0.07*** | -0.18*** | *0.74* |

*Note.* Intraclass correlation coefficients (ICCs) are shown on the diagonal (italicized). Between-person correlations are above the diagonal; within-person correlations are below the diagonal. † $p < .10$ , \* $p < .05$ , \*\* $p < .01$ , \*\*\* $p < .001$ .

### 3.2 Real-World Alcohol Cues and Momentary Craving

When all three real-world alcohol cues were modeled simultaneously (Table 3), higher-than-usual *in vivo* alcohol exposure was associated with higher momentary craving (*β* = 0.21, 95% CI [0.00, 0.42], *p* = .048). Similarly, being around usual drinking partners more than one’s typical level was independently associated with higher craving (*β* = 0.24, 95% CI [0.02, 0.45], *p* = .031). In contrast, usual drinking places were not significantly associated with craving. None of the corresponding between-person cue effects were statistically significant. All models adjusted for intervention phase, including the two interaction models (Tables 4 and 5), and post-intervention phase was consistently associated with lower momentary craving across models (*β* ranged from −0.23 to −0.27, *p* < .01).

**Table 3.** Multilevel Model Predicting the Effects of Real-World Alcohol Cues on Momentary Craving (N = 33; Observations = 2,369)

| Fixed Effects | Est. | SE | t | 95% CI |
| --- | --- | --- | --- | --- |
| Intercept | -1.52 | 2.29 | -0.67 | [-6.00, 2.96] |
| Age | 0.02 | 0.03 | 0.70 | [-0.03, 0.07] |
| Gender (Male = 1, Female = 0) | 0.31 | 0.67 | 0.46 | [-1.00, 1.61] |
| Education (Last Grade) | 0.19 | 0.25 | 0.74 | [-0.31, 0.68] |
| Ethnicity (Hispanic = 1, Others = 0) | 0.46 | 0.89 | 0.51 | [-1.28, 2.19] |
| Perceived Financial Status | 0.43 | 0.36 | 1.20 | [-0.27, 1.13] |
| AUDIT total (centered) | 0.10* | 0.04 | 2.39 | [0.02, 0.18] |
| Intervention (1 = Post, 0 = Pre) | -0.23** | 0.08 | -2.96 | [-0.39, -0.08] |
| Afternoon (vs. morning) | 0.39*** | 0.09 | 4.14 | [0.21, 0.58] |
| Evening (vs. morning) | 1.00*** | 0.09 | 11.47 | [0.83, 1.17] |
| Weekend (vs. weekday) | -0.02 | 0.08 | -0.19 | [-0.17, 0.14] |
| *In Vivo* Alcohol Exposure (BP) | 3.23 | 2.08 | 1.55 | [-0.86, 7.32] |
| *In Vivo* Alcohol Exposure (WP) | 0.21* | 0.11 | 1.98 | [0.00, 0.42] |
| People Usually Drink (BP) | 3.11 | 2.48 | 1.26 | [-1.74, 7.97] |
| People Usually Drink (WP) | 0.24* | 0.11 | 2.16 | [0.02, 0.45] |
| Place Usually Drink (BP) | -0.99 | 1.36 | -0.73 | [-3.65, 1.68] |
| Place Usually Drink (WP) | -0.03 | 0.09 | -0.33 | [-0.21, 0.15] |
| Random Effects | Variance |  |  |  |
| Intercept Variance (ID) | 2.43 |  |  |  |
| Residual Variance | 2.76 |  |  |  |

*Note.* BP = Between-person; WP = Within-person. Model includes a random intercept for participant. Time of day is coded as two dummy indicators (Afternoon and Evening) with Morning as the reference category. 95% CIs are Wald-based. $\dagger p < .10$ , $*p < .05$ , $**p < .01$ , $***p < .001$ . Models estimated using REML with Satterthwaite's degrees of freedom.

**Table 4.** Multilevel Model Predicting the Interactive Effects of Positive Affect and Real-World Alcohol Cues on Momentary Craving (N = 33; Observations = 2,369)

| Fixed Effects | Est. | SE | t | 95% CI |
| --- | --- | --- | --- | --- |
| Intercept | -1.40 | 2.01 | -0.70 | [-5.33, 2.54] |
| Age | 0.02 | 0.02 | 1.01 | [-0.02, 0.07] |
| Gender (Male = 1, Female = 0) | 0.20 | 0.58 | 0.35 | [-0.94, 1.34] |
| Education (Last Grade) | 0.13 | 0.22 | 0.60 | [-0.30, 0.56] |
| Ethnicity (Hispanic = 1, Others = 0) | 1.11 | 0.78 | 1.42 | [-0.42, 2.63] |
| Perceived Financial Status | 0.30 | 0.33 | 0.93 | [-0.34, 0.94] |
| AUDIT total (centered) | 0.07† | 0.04 | 2.03 | [0.00, 0.14] |
| Intervention (1 = Post, 0 = Pre) | -0.26*** | 0.08 | -3.35 | [-0.41, -0.11] |
| Afternoon (vs. morning) | 0.35*** | 0.09 | 3.82 | [0.17, 0.53] |
| Evening (vs. morning) | 0.94*** | 0.08 | 11.10 | [0.78, 1.11] |
| Weekend (vs. weekday) | -0.03 | 0.08 | -0.36 | [-0.18, 0.12] |
| *In Vivo* Alcohol Exposure (BP) | 0.69 | 1.87 | 0.37 | [-2.97, 4.35] |
| *In Vivo* Alcohol Exposure (WP) | 0.19† | 0.11 | 1.81 | [-0.02, 0.40] |
| People Usually Drink (BP) | 0.07 | 2.29 | 0.03 | [-4.41, 4.56] |
| People Usually Drink (WP) | 0.25* | 0.11 | 2.23 | [0.03, 0.47] |
| Place Usually Drink (BP) | 0.38 | 1.18 | 0.33 | [-1.92, 2.68] |
| Place Usually Drink (WP) | -0.11 | 0.20 | -0.56 | [-0.50, 0.28] |
| Positive Affect (BP) | -0.06 | 0.06 | -1.08 | [-0.17, 0.05] |
| Positive Affect (WP) | 0.04 | 0.02 | 1.67 | [-0.01, 0.08] |
| PA (WP) × *In Vivo* Alcohol Exposure (WP) | 0.08* | 0.04 | 2.14 | [0.01, 0.16] |
| PA (WP) × People Usually Drink (WP) | -0.03 | 0.04 | -0.72 | [-0.10, 0.05] |
| PA (WP) × Place Usually Drink (WP) | -0.05 | 0.03 | -1.54 | [-0.11, 0.01] |
| *Random Effects* | *Variance* |  |  |  |
| Intercept Variance (ID) | 2.68 |  |  |  |
| Positive Affect (WP) Slope Variance | 0.01 |  |  |  |
| Place Usually Drink (WP) Slope Variance | 0.85 |  |  |  |
| Residual Variance | 2.50 |  |  |  |

*Note.* BP = Between-person; WP = Within-person; PA = Positive Affect (happy + energetic). Random-effects structure: random intercept plus random slopes for PA (WP) and Place usually drink (WP), selected via systematic likelihood-ratio testing. 95% CIs are Wald-based. † $p < .10$ , \* $p < .05$ , \*\* $p < .01$ , \*\*\* $p < .001$ . Models estimated using REML with Satterthwaite's degrees of freedom.

**Table 5.** Multilevel Model Predicting the Interactive Effects of Negative Affect and Real-World Alcohol Cues on Momentary Craving (N = 33; Observations = 2,369)

| Fixed Effects | Est. | SE | t | 95% CI |
| --- | --- | --- | --- | --- |
| Intercept | -1.17 | 1.47 | -0.80 | [-4.05, 1.71] |
| Age | 0.03† | 0.02 | 1.86 | [0.00, 0.06] |
| Gender (Male = 1, Female = 0) | 0.20 | 0.42 | 0.48 | [-0.62, 1.02] |
| Education (Last Grade) | 0.04 | 0.16 | 0.25 | [-0.27, 0.35] |
| Ethnicity (Hispanic = 1, Others = 0) | -0.04 | 0.57 | -0.06 | [-1.15, 1.08] |
| Perceived Financial Status | 0.36 | 0.22 | 1.59 | [-0.08, 0.79] |
| AUDIT total (centered) | 0.09** | 0.03 | 3.31 | [0.04, 0.14] |
| Intervention (1 = Post, 0 = Pre) | -0.27*** | 0.08 | -3.43 | [-0.42, -0.11] |
| Afternoon (vs. morning) | 0.37*** | 0.09 | 4.04 | [0.19, 0.54] |
| Evening (vs. morning) | 0.91*** | 0.08 | 10.91 | [0.75, 1.08] |
| Weekend (vs. weekday) | -0.01 | 0.08 | -0.08 | [-0.15, 0.14] |
| *In Vivo* Alcohol Exposure (BP) | 0.80 | 1.33 | 0.60 | [-1.82, 3.41] |
| *In Vivo* Alcohol Exposure (WP) | 0.22* | 0.11 | 2.05 | [0.01, 0.42] |
| People Usually Drink (BP) | -0.14 | 1.58 | -0.09 | [-3.23, 2.95] |
| People Usually Drink (WP) | 0.29** | 0.11 | 2.61 | [0.07, 0.51] |
| Place Usually Drink (BP) | -0.64 | 0.88 | -0.73 | [-2.36, 1.08] |
| Place Usually Drink (WP) | -0.19 | 0.22 | -0.86 | [-0.63, 0.25] |
| Negative Affect (BP) | 0.22*** | 0.04 | 5.37 | [0.14, 0.30] |
| Negative Affect (WP) | 0.06 | 0.04 | 1.53 | [-0.02, 0.14] |
| NA (WP) × *In Vivo* Alcohol Exposure (WP) | 0.01 | 0.03 | 0.15 | [-0.06, 0.07] |
| NA (WP) × People Usually Drink (WP) | -0.01 | 0.04 | -0.24 | [-0.08, 0.06] |
| NA (WP) × Place Usually Drink (WP) | 0.06* | 0.03 | 2.09 | [0.00, 0.12] |
| Random Effects | Variance |  |  |  |
| Intercept Variance (ID) | 1.49 |  |  |  |
| Negative Affect (WP) Slope Variance | 0.04 |  |  |  |
| Place Usually Drink (WP) Slope Variance | 1.16 |  |  |  |
| Residual Variance | 2.46 |  |  |  |

*Note.* BP = Between-person; WP = Within-person; NA = Negative Affect (anxious + stressed). Random-effects structure: random intercept plus random slopes for NA (WP) and Place usually drink (WP), selected via systematic likelihood-ratio testing. 95% CIs are Wald-based. † $p < .10$ , \* $p < .05$ , \*\* $p < .01$ , \*\*\* $p < .001$ . Models estimated using REML with Satterthwaite's degrees of freedom.

### 3.3 Affect Moderated Cue Reactivity

#### 3.3.1 Role of PA

PA significantly moderated the within-person association between *in vivo* alcohol exposure and momentary craving (*β* = 0.08, 95% CI [0.01, 0.16], *p* = .032; Table 4). As shown in Figure 1, *in vivo* alcohol exposure was associated with stronger craving at moments when participants reported higher-than-usual PA. PA did not moderate the effects of usual drinking partners or usual drinking places on craving.

**Figure 1.**
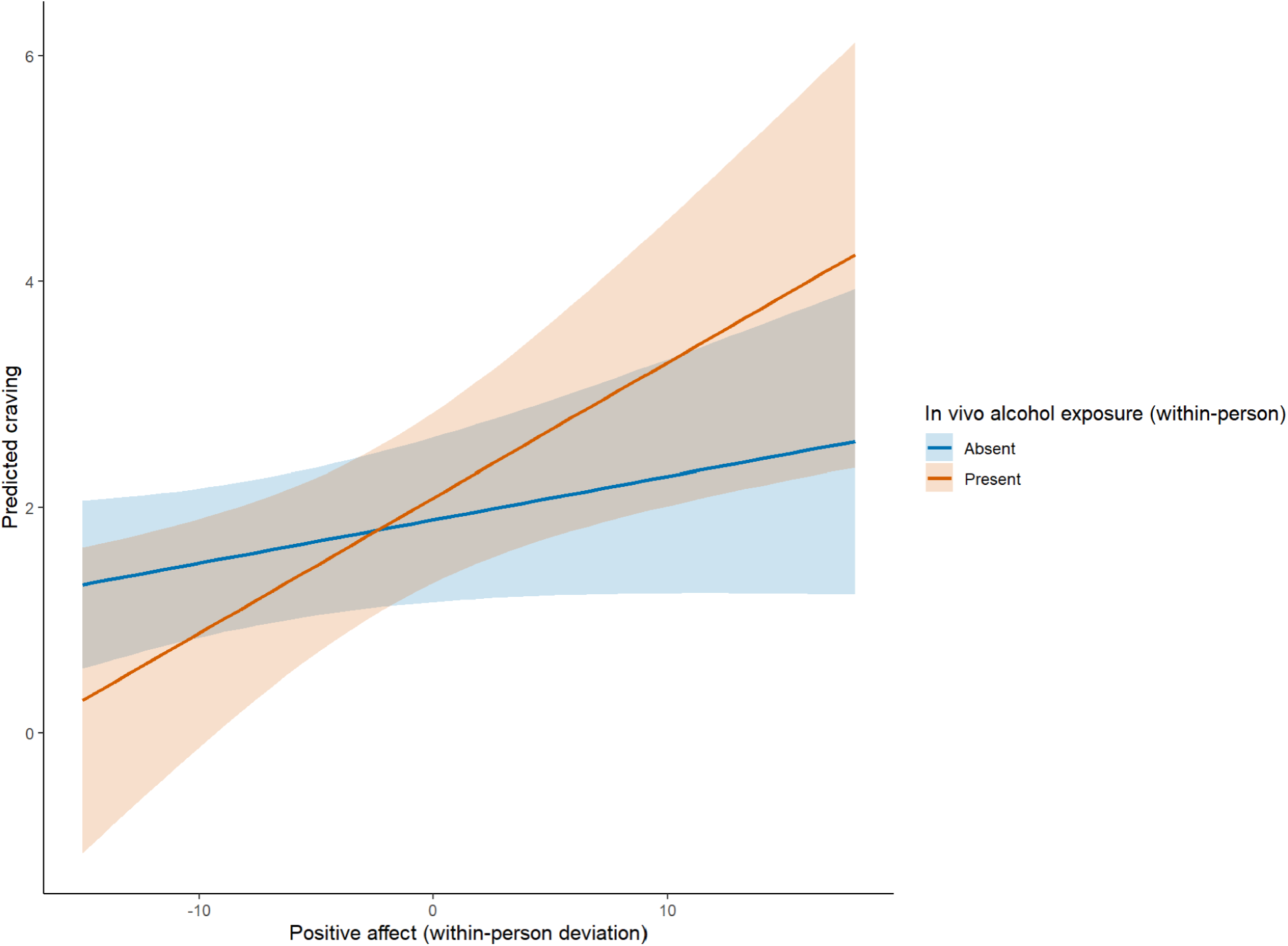
Within-person Positive Affect × *In Vivo* Alcohol Exposure on Momentary Craving *Note.* Predicted craving as a function of within-person positive affect and within-person *in vivo* alcohol exposure (0 = at participant’s typical exposure rate, 1 = elevated above typical). Shaded bands represent 95% confidence intervals.

#### 3.3.2. Role of NA

NA significantly moderated the within-person association between being in usual drinking places and momentary craving (*β* = 0.06, 95% CI [0.00, 0.12], *p* = .036; Table 5). As shown in Figure 2, being in usual drinking places was associated with stronger craving at moments when participants reported higher-than-usual NA. NA did not moderate the associations between *in vivo* alcohol exposure or being around usual drinking partners on craving.

**Figure 2.**
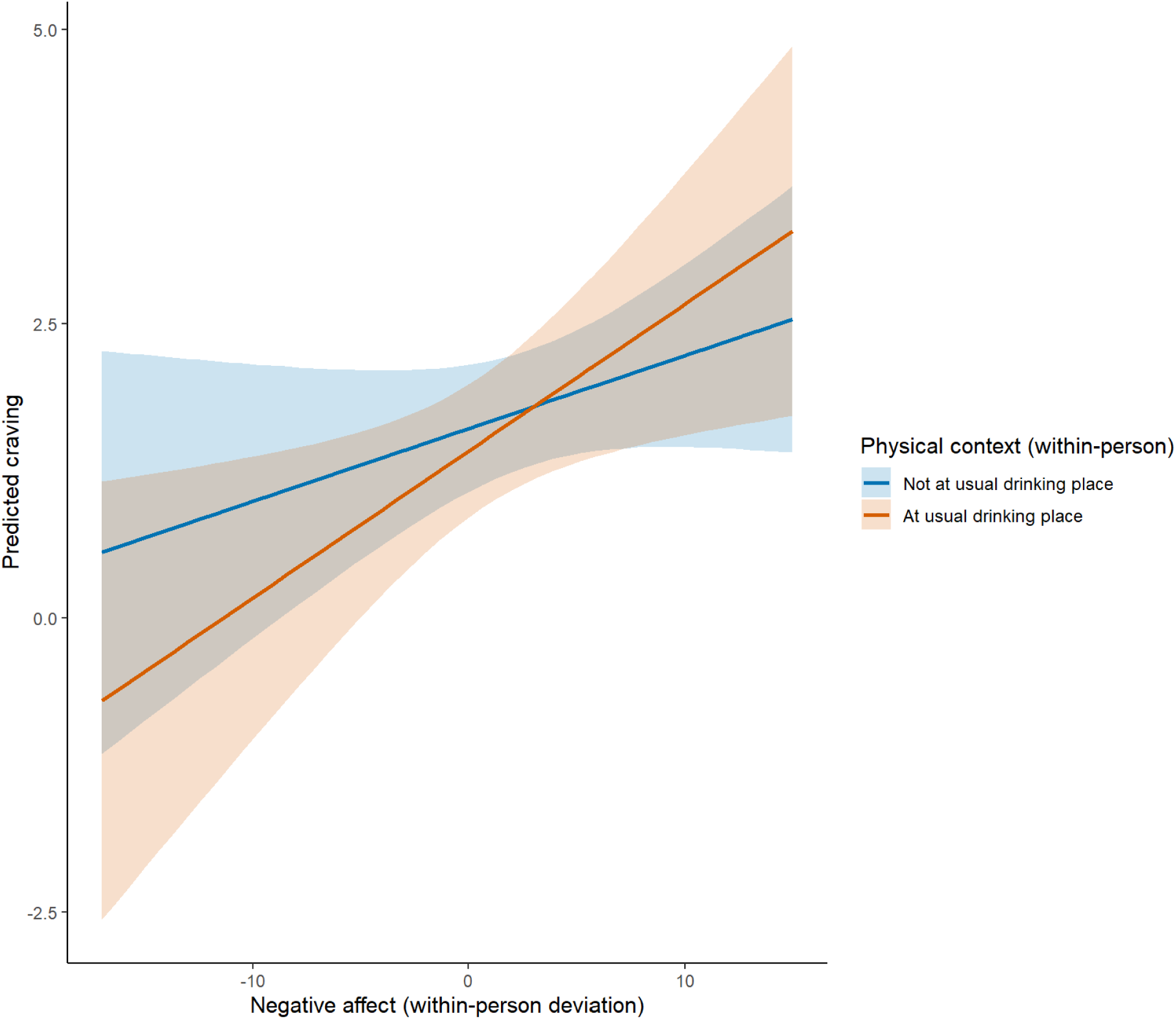
Within-person Negative Affect × Usual Drinking Place on Momentary Craving *Note.* Predicted craving as a function of within-person negative affect and within-person being at usual drinking place (0 = at participant’s typical exposure rate, 1 = elevated above typical). Shaded bands represent 95% confidence intervals.

## 4. Discussion

The present study used 28 days of smartphone-based EMA data from adults with AUD enrolled in an alcohol intervention study to examine how real-world alcohol cues and momentary affect jointly shape craving in daily life. Key findings revealed significant interaction effects, demonstrating that the within-person association between *in vivo* alcohol cues and momentary craving was stronger during moments when PA was higher-than-usual. Additionally, the within-person association between being in usual drinking places and momentary craving was stronger during moments when NA was higher-than-usual. These cue–affect associations were evident after adjusting for intervention phase, which was itself associated with lower average craving. Together, these findings can inform a layered approach to personalized intervention. Aggregated person-level cue–affect patterns could enhance clinician-delivered motivational interviewing by helping participants identify their own high-risk craving contexts. At the same time, the present findings suggest that low-frequency clinician-delivered feedback addresses average craving levels while momentary risk in specific cue–affect contexts may require a different intervention strategy. JITAIs may therefore be needed to address these high-risk moments more directly through real-time coping, affect-regulation, or reward-regulation support.

### 4.1 Real-World Alcohol Cues and Momentary Craving

The independent within-person associations of *in vivo* alcohol cues and personalized social cues with momentary craving are consistent with incentive sensitization and conditioning frameworks. *In vivo* alcohol exposure may operate as an immediate and perceptually salient cue that activates alcohol-related “wanting” through bottom-up incentive-salience processes (Berridge and Robinson, 2016; Cofresí et al., 2019; Robinson and Berridge, 2025). Although prior EMA studies have linked *in vivo* alcohol cues to craving primarily in adolescent and emerging-adult samples (Kohen et al., 2023; Treloar Padovano and Miranda, 2021; Wycoff et al., 2023), the present findings extend this evidence to treatment-motivated adults with AUD and show that *in vivo* alcohol exposure remains associated with craving even when personalized social and place-based cues are modeled simultaneously.

The independent association of personalized social cues with craving points to a related but distinct pathway. Participants reported whether they were around people with whom they usually drink, rather than whether they were around others in general. This item therefore captured a person-specific social cue that may reflect learned associations between particular interpersonal contexts and alcohol use. Its independent association with craving suggests personally meaningful social conditioning, whereby specific people become signals for drinking through repeated co-occurrence, making particular interpersonal contexts uniquely craving-relevant (Acuff et al., 2024; Fairlie et al., 2025; Kuerbis et al., 2020; Meisel et al., 2023).

The null within-person association between usual drinking places and craving should not be interpreted as evidence that place cues are unimportant. The place cue had the highest occurrence, with participants reporting being in a usual drinking place at 41% of moments, suggesting that it may have captured a common exposure rather than an acute craving-relevant event. In addition, when *in vivo*, social, and place cues are modeled together, the unique effect of place exposure may be reduced. While prior EMA work has shown some evidence that AUD severity strengthens context-related craving risk, these studies have not typically tested different cues within the same model, leaving unclear whether place cues uniquely predict craving (Fatseas et al., 2015; Kuerbis et al., 2020).

### 4.2 PA Strengthens In Vivo Cue Reactivity

High-arousal PA significantly amplified the association between *in vivo* alcohol exposure and momentary craving. This finding is consistent with positive-reinforcement and enhancement models of alcohol use, in which drinking is motivated by the desire to extend, intensify, or capitalize on pleasant and activated states rather than only to reduce distress (Cooper, 1994; Cooper et al., 1995). Recent experience sampling method evidence among women with AUD similarly found that within-person increases in PA were curvilinearly associated with momentary craving and predicted both non-heavy drinking and binge drinking, with PA emerging as a stronger momentary driver of craving than NA in this clinical group (Leenaerts et al., 2025). This pattern is also consistent with broader daily-life evidence suggesting that drinking may be more likely on high-positive-affect days than high-negative-affect days, with most evidence for college-age, nonclinical samples (Dora et al., 2023). The present findings extend this literature to an AUD intervention study and cue reactivity by showing that pleasant, activated states are not necessarily protective against craving, rather may heighten reactivity to *in vivo* cues among adults with AUD.

### 4.3 NA Strengthens Place Cue Reactivity

Within-person elevations in NA showed evidence of strengthening the association between being in usual drinking places and momentary craving. This pattern is theoretically coherent. Self-medication and affective-processing models suggest that distress states can become powerful craving triggers when repeatedly paired with alcohol’s relief-producing effects (Baker et al., 2004; Khantzian, 1997). When distress occurs in environments strongly associated with drinking, internal affective cues and external place cues may converge to heighten craving. This interpretation aligns with relapse models emphasizing the combined influence of affective distress and high-risk environments (Marlatt, 1996; Witkiewitz and Marlatt, 2004), as well as intensive longitudinal reviews suggesting that NA–craving associations in treatment-seeking samples may be better understood through interaction frameworks than main effects alone (Votaw et al., 2022). Neurobiological evidence further supports this possibility. In adults with AUD, acute psychosocial stress has been shown to sensitize cue-elicited insular activation, and laboratory stress reactivity prospectively predicts craving and alcohol use during ambulatory follow-up (Bach et al., 2024; Zaiser et al., 2025).

### 4.4 Clinical Implications for Just-in-Time Adaptive Interventions

These findings have direct implications for developing personalized-feedback interventions and JITAIs for AUD. In the present alcohol intervention, participants received a clinician-delivered personalized report that summarized aggregated EMA reports of drinking precursors to bolster the motivational interviewing intervention, and the post-intervention phase was associated with lower average craving. At the same time, the cue–affect associations observed after adjustment for intervention phase suggest that low-frequency personalized feedback may not fully address momentary risk as it unfolds in real time. JITAIs may therefore complement clinician-delivered feedback by delivering support during discrete high-risk moments. For example, moments when an individual is exposed to *in vivo* cues while experiencing higher-than-usual PA may call for strategies that target reward anticipation and approach motivation, such as brief mindfulness prompts, or substitution with alternative rewarding activities. In contrast, moments when an individual experiences higher-than-usual NA in usual drinking places may call for distress-tolerance or emotion-regulation strategies. This type of tailoring aligns with emerging evidence that brief, contextually tailored messages can improve proximal intervention targets, such as drinking self-efficacy, shortly after delivery (Dauber et al., 2025). However, systematic reviews suggest that the JITAI evidence base for substance use remains nascent, with many studies focused on adolescents and young adults rather than adults with established drinking patterns or AUD (Koike et al., 2025). Building on these findings, future JITAIs for AUD could test whether moments involving *in vivo* cues and elevated PA, or usual drinking places and elevated NA, are optimal points for delivering tailored support aimed at reducing craving and subsequent drinking risk.

### 4.5 Limitations and Future Directions

The current study has several limitations. First, the modest person-level sample size (N = 33) may have limited power to detect smaller cue-by-affect interactions and constrains the generalizability of the findings. Future studies should replicate these findings in larger samples of adults with AUD. Second, the data were observational, and the interaction effects were estimated concurrently within the same EMA prompt rather than prospectively across time. Future studies with larger samples and more observations should use dynamic multilevel models to test whether specific cue–affect combinations prospectively predict subsequent increases in craving within and across days. Third, alcohol cues, affect, and craving were self-reported, which may introduce reporting bias, and random EMA prompts may miss brief cue-reactivity episodes occurring between assessments. Future studies should integrate passive sensing approaches to capture contextual risk more continuously. Recent work suggests that passive sensing is acceptable to adults with AUD in early recovery, and wearable transdermal alcohol biosensors paired with machine-learning decision rules may support real-time, context-aware JITAI delivery (Fairbairn et al., 2024; Wang et al., 2024; Wyant et al., 2023). Finally, the PA and NA composites included only two items each and focused on high-arousal affective states. More granular affect measurement is needed to distinguish high- and low-arousal forms of PA and NA and to test whether arousal, valence, or their combination differentially shapes cue reactivity in AUD.

## 5. Conclusions

The current study suggests that real-world alcohol cues and affective states may jointly shape momentary alcohol craving among treatment-motivated adults with AUD. Specifically, high-arousal PA strengthened the association between *in vivo* alcohol exposure and craving, whereas high-arousal NA strengthened the association between usual drinking places and craving. These cue–affect associations were evident after adjustment for intervention phase, which was associated with lower average craving, suggesting that clinician-delivered motivational interviewing may reduce overall craving while real-time cue–affect risk may require complementary momentary intervention strategies. By identifying moments when craving may intensify, such as exposure to *in vivo* alcohol cues during elevated PA or usual drinking places during elevated NA, this work may inform personalized JITAIs that deliver coping, reward-regulation, or distress-tolerance support during heightened risk. Future research using larger samples, lagged designs, and passive sensing methods will be needed to refine these models and evaluate whether cue–affect-informed interventions reduce craving and subsequent drinking among individuals with AUD.

## Author Contributions (CRediT)

**Abhishek Aggarwal:** Conceptualization, Formal analysis, Methodology, Visualization, Writing – original draft, Writing – review & editing. **Peter M. Monti:** Conceptualization, Funding acquisition, Supervision, Writing – review & editing. **Kittichai Promrat:** Investigation, Resources, Writing – review & editing. **Molly Magill:** Writing – review & editing. **Jessica L. Mellinger:** Writing – review & editing. **Hayley Treloar Padovano:** Conceptualization, Data curation, Funding acquisition, Investigation, Methodology, Project administration, Supervision, Writing – review & editing.

## Data Availability Statement

Data are not publicly available due to restrictions arising from the informed-consent agreement and our IRB’s policy regarding the privacy of participants with alcohol use disorder. De-identified data may be made available to qualified investigators following execution of a Data Use Agreement and approval by the Brown University IRB. Requests should be directed to the corresponding author.

